# Taking Account of Asymptomatic Infections – A Modeling Study on the COVID-19 Outbreak on the Diamond Princess Cruise Ship

**DOI:** 10.1101/2020.04.22.20074286

**Authors:** Li-Shan Huang, Li Li, Lucia Dunn, Mai He

## Abstract

**Background:** The COVID-19 outbreak on the Diamond Princess (DP) cruise ship provided empirical data to study the transmission potential of COVID-19 under quarantine with the presence of asymptomatic cases.

**Methods:** We studied the changes in *R*_0_ on the DP from January 21 to February 19, 2020 based on chain-binomial models under two scenarios: no quarantine assuming a random mixing condition, and quarantine of passengers in cabins — passengers may get infected either by an infectious case in a shared cabin or by asymptomatic crew who continued to work.

**Results:** Estimates of *R*_0_ at the beginning of the epidemic were 3.27 (95% CI, 3.02-3.54) and 3.78 (95% CI, 3.49-4.09) respectively for serial intervals of 5 and 6 days; and when quarantine started, with the reported asymptomatic ratio 0.505, *R*_0_ rose to 4.18 (95%CI, 3.86-4.52) and 4.73 (95%CI, 4.37-5.12) respectively for passengers who might be exposed to the virus due to contacts with asymptomatic crew. The overall *R*_0_ for both crew and passengers was decreased to 2.55 (95%CI, 2.36-2.76) and 2.90 (95% CI, 2.67-3.13). Results show that the higher the asymptomatic ratio is, the more infectious contacts would happen.

**Conclusions:** We find evidence to support a US CDC report that “a high proportion of asymptomatic infections could partially explain the high attack rate among cruise ship passengers and crew.” Our study suggests that the effects of quarantine may be limited if the asymptomatic ratio is high, implying that a combination of preventive measures is needed to stop the spread of virus.

## Introduction

The COVID-19 outbreak has developed into an international public health emergency [1]. The reproductive number (*R*_0_) of COVID-19 is a key piece of information for understanding an epidemic. Current intervention methods focus on quarantine methods with either mitigation or suppression strategies aimed at reducing the reproduction number *R*_0_ and flattening the curve [2]. Asymptomatic infectious cases are less likely to seek medical care or to be tested and quarantined, contributing to the infectious potential of a respiratory virus [3,4]. Clinical findings have suggested that the viral load in asymptomatic patients is similar to that in symptomatic patients [5]. Evidence suggests that these asymptomatic patients can infect others before they manifest any symptoms [6-8]. In an early study [9] of cases in Wuhan, China, 200 individuals out of 240 (83%) reported no exposure to an individual with respiratory symptoms, which suggests pre-symptomatic/asymptomatic infection is common [10]. The Diamond Princess (DP) data with reported asymptomatic cases [11-13] may be considered as an “accidental” trial in an isolated environment. Based on the DP data [11-13], we estimate the *R*_0_ as a function of time, and our approaches take explicit account of possibly infectious contacts between quarantined passengers in cabins and asymptomatic crew, which has not been explored in the literature. A US CDC report states that “a high proportion of asymptomatic infections could partially explain the high attack rate among cruise ship passengers and crew [14].”

The DP [11-13], with 3,711 people (2,666 passengers and 1,045 crew members) on board as of February 5, 2020, was found to have an outbreak of COVID-19 from one traceable passenger from Hong Kong. This passenger became symptomatic on January 23 and disembarked on January 25 in Hong Kong. On February 1, six days after leaving the ship, he tested positive for SARS-CoV-2 at a Hong Kong hospital. Japanese authorities were informed about this test result. On February 4, the authorities announced positive test results for SARS-CoV-2 for another ten people on board. The ship was quarantined by the Japanese Ministry of Health, Labour and Welfare for what was expected to be a 14-day period, off the Port of Yokohama [12]. Initially, passengers were quarantined in their cabins while the crew continued to work. Only symptomatic cases and close contacts were tested for COVID-19 and PCR-confirmed positive passengers were removed and isolated in Japanese hospitals. As reported, phased attempts were made to test all passengers including asymptomatic cases starting on February 11. As of February 20, 619 cases had been confirmed (16.7 % of the population on board), including 82 crew and 537 passengers [11]; and 50.5% of the COVID-19 cases on the DP were asymptomatic [13], while an estimated proportion 17.9% (95% credible interval: 15.5–20.2%) never demonstrated any symptoms, based on a Bayesian modeling approach [13]. Overall, 712 (19.2%) of the crew and passengers tested positive [14].

The *R*_0_ of COVID-19 on DP has been estimated previously [15]; this research identified the *R*_0_ as 14.8 initially and then declining to a stable 1.78 after the quarantine and removal interventions, assuming a 70% reduction in contact rate. That research does not take account of asymptomatic cases. Other researchers using the DP data up to February 16 have estimated the median *R*_0_ as 2.28 [16]. They found *R*_0_ remained high despite quarantine measures, while concluding that estimating *R*_0_ was challenging due to the difficulty in identifying the exact number of infected cases. The *R*_0_ values have important implications for predicting the effects of interventions. The threshold for combined vaccine efficacy and herd immunity needed for disease extinction is 1-1/*R*_0_. At *R*_0_=2, the threshold is 50%, while at *R*_0_=4, this threshold increases to 75%.

We investigated the changes in *R*_0_ for COVID-19 on the DP from January 21 to February 19 with a chain-binomial model at different times under two scenarios: no quarantine assuming a random mixing condition, and quarantine of passengers in cabins — passengers may get infected either by an infectious case in a shared cabin or by asymptomatic crew who continued to work. This work adds to the growing knowledge gained from the DP data in that we estimate *R*_0_ by (1) mimicking the quarantine conditions in practice, and (2) taking explicit account of the presence of the asymptomatic crew and phased removal of infectious cases.

## Materials and Methods

### Data

We collected publicly available data on the outbreak on the DP from January 21 to February 19 [11-13]. We set January 21 as day 1, since January 20 was the start date (day 0) of the cruise. February 19 (day 30) was the date that most passengers were allowed to leave the ship. For those dates that Y, the number of new COVID-19 cases, was not reported, linear interpolation was used. As an example, there were 67 new cases on February 15, but no data were reported on February 14. After linear interpolation, *Y* on a daily basis became 33 and 34 for February 14 and 15 respectively. Based on the documented onset dates [11], there were 34 cases with onset dates before February 6, and we further adjusted the number of confirmed cases on February 3, 6, and 7, from 10, 10, and 41 cases to 17, 17, and 27 cases respectively. We chose serial intervals τ of 5 and 6 days as these are factors of 30 and are close to 7.5 days (95% CI 5.3-19) [9] and 4 days [17,18]. Then daily data were aggregated into 5- and 6-day intervals.

### Statistical Analysis

The chain-binomial model originally proposed in [19], belongs to the broader class of stochastic discrete-time SIR models [20]. The model assumes that an epidemic is formed from a succession of generations of infectious individuals from a binomial distribution. For the DP data, the initial population size is *N_t_*_=0_ = 3711, where time *t* is the duration measured in units of the serial interval. To model the dynamics on the ship for the case τ = 6, we make the following assumptions, which are stated in the order of time.

a. From January 21 to 26 (*t* = 1, the first serial interval), infection contacts happened at random following the random mixing assumption. Let *I_t_* be the number of persons infected at time *t*. Then *I_t_*_=_*_1_* is a binomial random variable *B*(*N_t_* =0*, p_1_*) with binomial transmission probability *p*_1_ = 1 - exp(- β × *I_t_*_=0_*/N_t_*=0), where β is the transmission rate and *I_t_*_=0 = 1_ (the first case who disembarked on January 25). As in the SIR model, the probability that a subject escapes infectious contact is assumed to be exp(- β × *I_t_*_=0_/*N*_*t*=0_).
b. From January 27 to February 1 (*t* = 2), infection contacts again followed the random mixing assumption. Hence *I_t_*_=2_ is a binomial random variable *B*(*N_t_* =*1, p*_2_) with *p*_2_ = 1 - exp(- β × *I_t_*_=1_*/N_t_*_=0_), and the number of persons at risk of infection is *N_t_* _=1_ = 3711 - *I_t_*_=1_.
c. For the period February 2 to 7 (*t* = 3), *I_t_*_=3_ is a binomial random variable *B*(*N_t_* _=2_, *p*_3_) with *N_t_* _=2_ = *N_t_* _=1_ - *I_t_*_=2_ and *p*_3_ = 1 - exp(- β × (*I_t_*_=_1 + *I_t_*_=_*2*)*/N_t_* _=0_). As the quarantine started on February 5 and confirmed cases were removed, the number of persons infected, removed, and at risk of infection at the end of the *t*=*3* period were *I_t_*_=3_, (*I_t_*_=1_ + *I_t_*_=2_), and *N_t_* _=3_ = *N_t_* _=2_ - *I_t_*_=3_ respectively.
d. For *t* = 4 and *t* = 5 during the quarantine of passengers, *N_t_* = *N_t_*_-1_ *- I_t_*, and infectious cases were removed. We further make the following assumptions. (i) Of all infected cases, 86.8%(=537/619) were passengers and 13.2% (=82/619) crew [11]. We use these proportions to calculate the number of infected persons in each group, *Ip_t_* and *Ic_t_*, respectively, and *I_t_* = *Ip_t_* + *Ic_t_*. This assumption is imposed since there is no public data available for the time course of *Ip_t_* and *Ic_t_*. (ii) Crew members continued to work unless showing symptoms; hence the binomial transmission probability of crew remained the same as 1 – exp(– β × *I_t_*_-1_*/N_t_*_-1_), *t* = 4 and 5. In other words, crew were randomly mixing in the population on board. (iii) Passengers stayed in cabins most of the time. Assume that among infected passengers *Ip_t_*, the proportion of infections that occurred in cabins is *r_p_*, and that the average occupancy per cabin is 2. For those *Ip_t_*_-1_ cases, *t* = 4, 5, the binomial transmission probability to infect *Ip_t_ × r_p_* passengers in cabins is 1 - exp(- β/2). In [11], *r_p_* = 0.2 (= 23/115), while we assume *r_p_* = 0.2 and 0.3 when τ = 6. For this assumption, *r_p_* ≤ *Ip_t-1_/Ip_t_*, and thus *r_p_* cannot be made arbitrarily large. (iv) The other (1 - *r_p_*) proportion of infected passengers’ cases was possibly due to asymptomatic crew members who continued to perform service [11], and their binomial transmission probability is assumed to be *p_t_* = 1 - exp(- β × *aratio × Ic_t_*_-1_/*C_t_*_-0_, where *aratio* is the asymptomatic ratio and *C_t_*_-1_ is the number of crew members on board at time *t* - 1. That is to say, these passengers were randomly mixing in the crew population with possible infectious contact with asymptomatic crew. In our calculations, *aratio* = 0.4, 0.465 [14], 0.505 [13], and 0.6.

Assumptions (a)-(c) correspond to no quarantine assuming a random mixing condition, and assumption (d) to quarantine of passengers in cabins in which passengers may either get infected (d)(iii) by an infectious case in a shared cabin or (d)(iv) by asymptomatic crew who continued to work. The maximum likelihood (ML) approach was used to estimate β. We developed some R code [20] by modifying some R-functions in [21] for the chain-binomial model and the ML step was carried out using the R-package bbmle [22]. The associated R-code is provided in the eAppendix. For *t* = 4, 5, *R*_0_ is the number of persons (passengers or crew) at risk (at time *t*) times either (d)(iv) 1 - exp(- β × *aratio/ C_t_*_-1_) for passengers potentially infected by asymptomatic crew, or (d)(ii) 1 - exp(-β */N_t-1_*) for crew.

The calculation for the case τ = 5 is analogous: period January 21 to 25 follows (a); period January 26 to 30 follows (b); period January 31 to February 4 follows (c); and periods February 5 to 9, February 10 to 14, and February 15 to 19 follow (d), and *r_p_* = 0.2, which is the maximum value given the constraint *r_p_* ≤ *Ip_t-1_/Ip_t_*.

## Results

For the DP COVID-19 outbreak, Table 1 gives the estimates of β and their 95% confidence intervals. Since β is the basic reproductive number *R*_0_ at the beginning of the epidemic (*t* = 1, 2, 3 when τ = 5, and *t* = 1, 2 when τ = 6), we observe from Table 1 that the estimated *R*_0_ for the initial period is greater than 3 in every one of the scenarios that we considered. In addition, given τ and *r_p_*, the estimated β increases as *aratio* decreases. Thus if the *aratio* is smaller than 40%, the estimated β would be larger than those in Table 1.

**Table 1:** Estimates of β and their 95% confidence intervals for the Diamond Princess COVID-19 outbreak data.

| asymptomatic<br>ratio (*aratio*) | $\beta = R_0$ at $t=1, 2$ when $\tau = 6$, at $t=1, 2, 3$ when $\tau = 5$; estimate (95% CI) | | |
| --- | --- | --- | --- |
| | $\tau = 6; r_p = 0.2$ | $\tau = 6; r_p = 0.3$ | $\tau = 5; r_p = 0.2$ |
| 40% | 3.94 (3.64, 4.27) | 4.41 (4.06, 4.78) | 3.41 (3.15, 3.69) |
| 46.50% | 3.84 (3.54, 4.16) | 4.28 (3.94, 4.64) | 3.33 (3.07, 3.60) |
| 50.50% | 3.78 (3.49, 4.09) | 4.20 (3.87, 4.55) | 3.27 (3.02, 3.54) |
| 60% | 3.64 (3.36, 3.94) | 4.03 (3.71, 4.36) | 3.16 (2.91, 3.42) |

When *aratio*=0*.5*0*5* [13], the estimated *R*_0_ as a function of *t* and its 95% CI are given in Table 2 and illustrated in Figure 1 for the case of *r_p_* = 0.2. We observe that when τ =6 and *r_p_*=0.2, the *R*_0_ for passengers in (d)(iv) is increased from 3.78 at *t*=3 to 4.73 and 4.39 at *t* = 4, 5, respectively, and the *R*_0_ is decreased to 1.06 and 1.05 respectively for crew in (d)(ii). This shows that *R*_0_ for some passengers increased from *t* = 3 to *t* = 4, 5 if they were in contact with asymptomatic crew members. On the other hand, the *R*_0_ for crew at *t* = 4, 5 is small and close to since infected passengers were removed and crew were exposed to fewer cases. With a higher *r_p_* = 0.3, the same τ = 6 and *aratio* = 0.505, the estimated β=4.20 (Table 1) is larger than the case with *r_p_* = 0.2, and the *R*_0_ for passengers in (d)(iv) is increased to 5.26 and 4.88 at *t* = 4 and 5 respectively (Table 2); and the *R*_0_ for crew in (d)(ii) is decreased to 1.18 and 1.17 respectively. For the case of τ = 5, *r_p_* = 0.2, *aratio* = 0.505, and estimated β = 3.27 (Table 1 and Figure 1), Table 2 shows that the *R*_0_ for passengers in (d)(iv) is again increased to 4.18, 4.08, and 3.74 at *t* = 4, 5, and 6 respectively, and the *R*_0_ for crew in (d)(ii) is below 1, 0.92, 0.92, and 0.91 respectively.

**Table 2:**
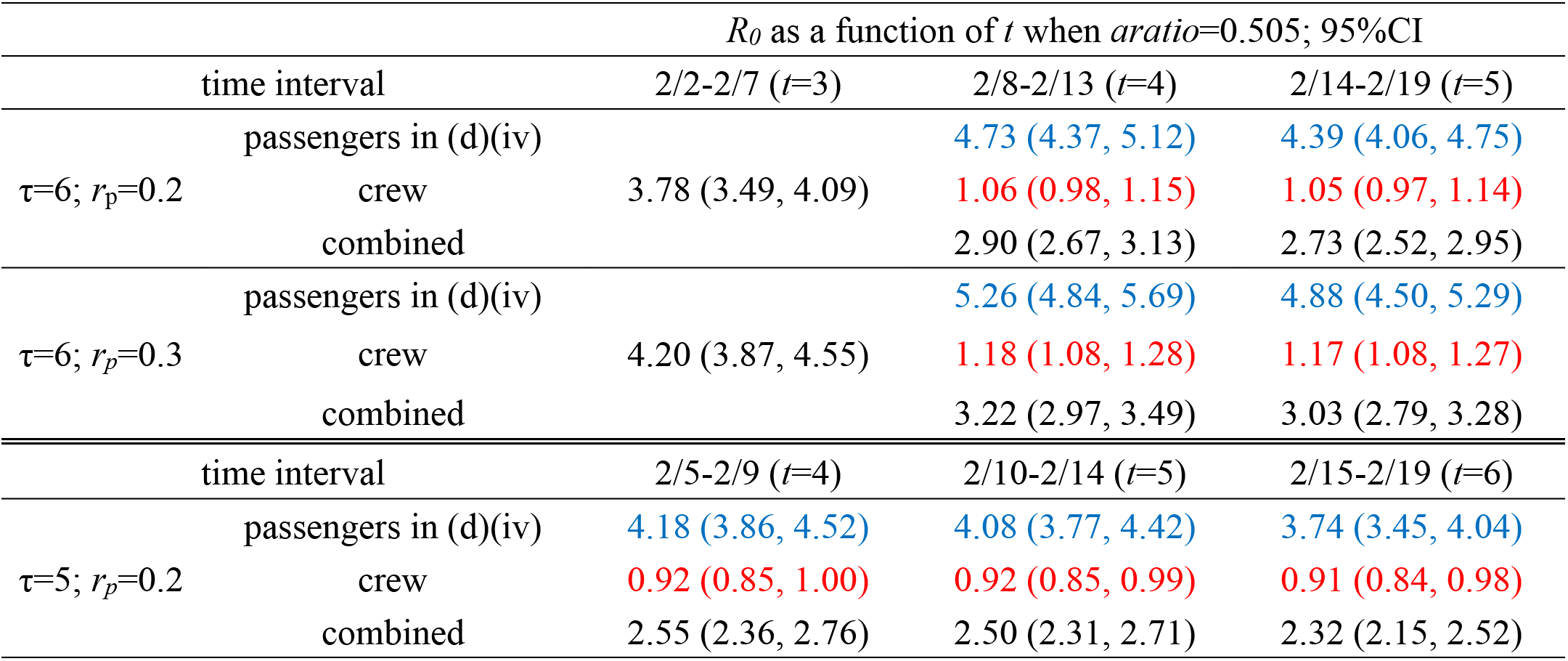
Estimates of *R*_0_ as a function of *t* and their 95% confidence intervals for the Diamond Princess COVID-19 outbreak.

**Figure 1:**
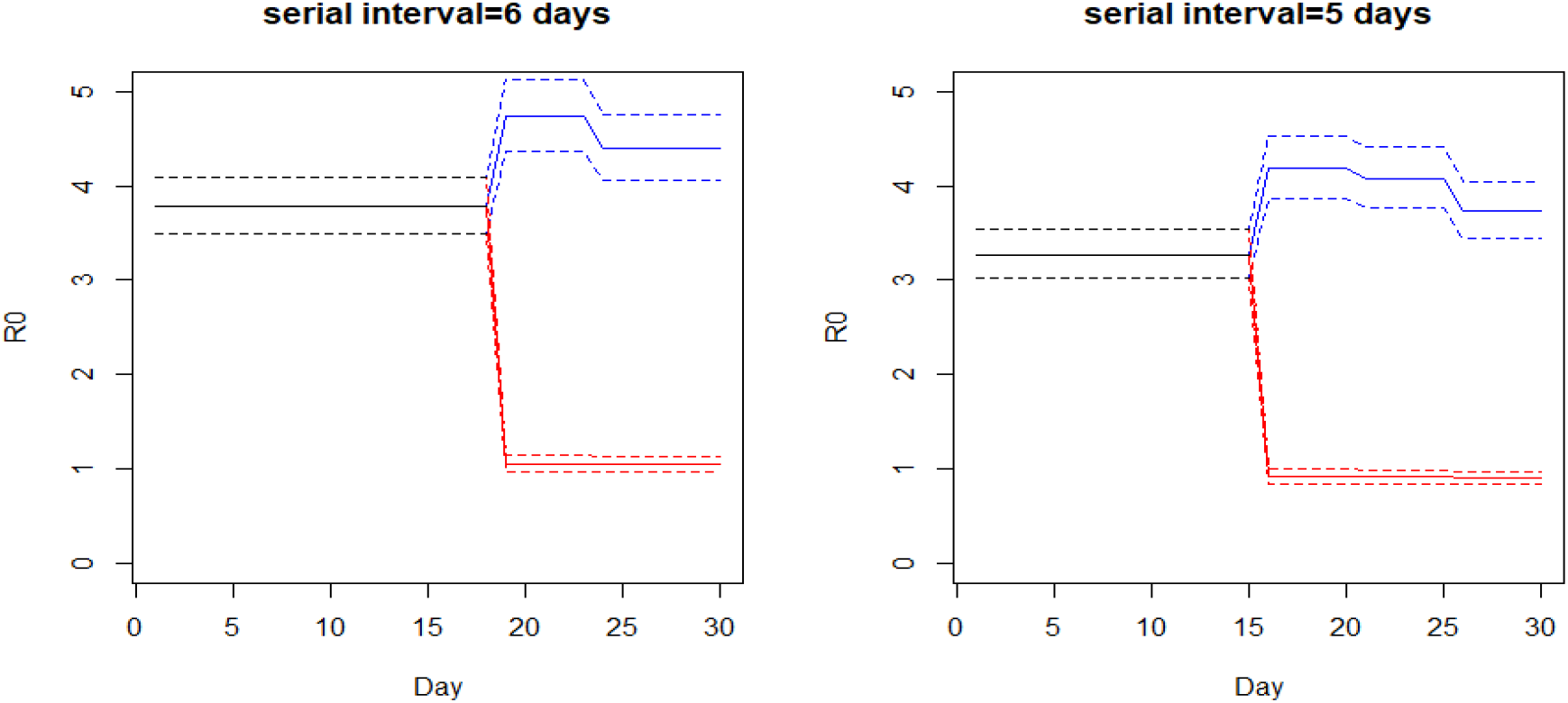
Time-dependent effective reproduction number *R*_0_ (solid lines) of COVID-19 on board the Diamond Princess ship January 21 (day 1) to February 19 (day 30) and their 95% confidence intervals, assuming τ = 5 and 6 days, *r_p_* = 0.2 and *aratio* = 0.505. Blue: *R*_0_ for passengers in contact with asymptomatic crew members. Red: *R*_0_ for crew members.

Other than those infections between passengers sharing the same cabin, the combined *R*_0_ for passengers and crew are also given in Table 2, 2.90 and 2.73 respectively for *t* = 4 and 5 when τ = 6, *r_p_* = 0.2, and *aratio* = 0.505, decreasing from the initial *R*_0_ = 3.78, which illustrates the limited effects of quarantine if asymptomatic cases were present. Similarly, when τ = 5, *r_p_* = 0.2, and *aratio* = 0.505, the combined *R*_0_ for passengers and crew is 2.55, 2.50, and 2.32 respectively for *t* = 4, 5 and 6. To understand the dynamics of no quarantine with a high *R*_0_, Figure 2 shows 100 stochastic simulations of the chain binomial model based on *N* = 3,700 and β = 3.78, assuming no quarantine, infected cases removed, τ = 6, and extrapolation to 90 days, with the observed epidemic (red line). It suggests that the quarantine on DP did prevent a more serious outbreak. If there was no quarantine, the cumulative number of cases at the end of 30 days has a mean 855 (SD = 439), and median 791 (IQR = 533), while the observed DP data of 621 cases [11] is at the 35th percentile. Among 99 of 100 simulations, the entire population is infected at the end of 54 days, and in the remaining one simulation, the entire population is immune, not infected at all.

**Figure 2:**
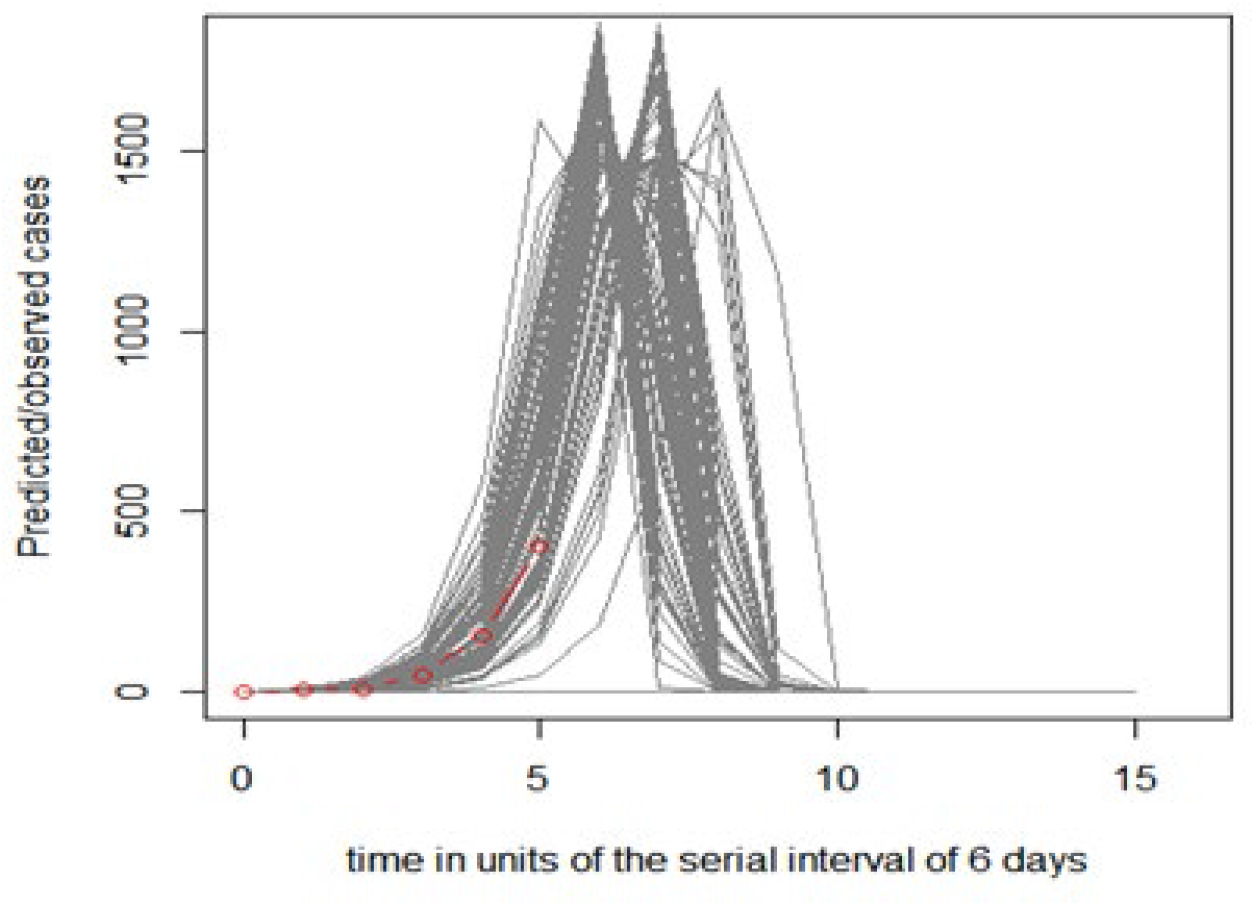
99 of 100 stochastic simulations of the chain binomial model based on *N* = 3700 and β = 3.78, assuming no quarantine, serial interval τ = 6 days, infected cases removed, and extrapolation to 90 days, with the observed epidemic (red line). For the remaining one simulation, the entire population is immune, not infected at all.

Let us explain mathematically why the *R*_0_ increased for passengers in contact with asymptomatic crew. Denote the number of passengers in assumption (d)(iv) as *Pa_t_*. To reduce their *R*_0_ to a number smaller than the initial β, a sufficient condition is *aratio*≤ *C_t_*_-1_*/ Pa_t_*, and the DP crew-passenger ratio at *t*=0 is 1045/2666=0.39 [12]. This suggests that as long as the *aratio* is ≥40%, *R*_0_ for passengers in (d)(iv) would remain high due to asymptomatic crew. The higher the *aratio* is, the more infectious contacts would happen. Thus, for a virus with a high *aratio*, the conventional quarantine procedure may not be effective to stop the spread of virus, highlighting the importance of early and effective surveillance.

We then derived a sufficient condition to achieve *R*_0_≤1 for both passengers and crew: β × *aratio* × *Pa_t_* ≤ *C_t-1_* and *C_t_ ≤ N_t −1_*/β. We attempt to interpret this condition as follows. For a β about 3, *C_t_* ≤ *N_t −1_*/β means that the number of people who continued to work during quarantine is less than one-third of the population, which is generally the case during quarantine. However, β × *aratio* × *Pa_t_* ≤ *C_t_*_-1_ may not be satisfied depending on the *aratio* value. When the *aratio* is close to 0, this condition is satisfied, but when *aratio* is high, the asymptomatic crew continue to spread the virus and passengers staying in cabins could not escape infectious contacts. This suggests that if the true *aratio* value is high, the current “stay-at-home” quarantine procedure may not be sufficient to reduce *R*_0_≤1 and to eliminate the virus completely.

## Discussion

The effects of pre/asymptomatic population on the spread of COVID-19 during quarantine has not yet been studied extensively. Clinical observations and lab tests have confirmed the existence of a pre/asymptomatic population infecting others [6-8]. It is not easy to give estimates of the size of this population, yet the DP outbreak provides useful real world data for this. 50.5% of passengers and crew members on the DP were asymptomatic and an estimated 17.9% of the infected individuals never demonstrated any symptoms [13].

In this study, the estimated *R*_0_ for the initial period (Table 1) are all greater than 3, consistent with most estimates of *R*_0_ reported earlier, showing that the COVID-19 virus is highly contagious [23,24]. The initial *R*_0_ is also comparable to those estimated in other non-cruise settings. The novelty of our approach has been to incorporate the asymptomatic ratio in the chain-binomial model to account for the possibly infectious contacts between quarantined passengers and asymptomatic crew. The results show that with a serial interval of 6 days, *R*_0_ is similar for *t* = 1-3, yet *R*_0_ for some passengers in assumption (d)(iv) is higher for *t* = 4, 5. The results show that the observed proportions of infections, 86.8%(=537/619) for passengers and 13.2% (=82/619) for crew [11], is possible and we find evidence to support a US CDC report that “a high proportion of asymptomatic infections could partially explain the high attack rate among cruise ship passengers and crew [14].”

Some research has suggested that the pre/asymptomatic population, “silent carriers,” are the main driving force behind this pandemic. A group [25] has estimated that the proportion of undocumented infections in China — including those who experience mild, limited or no symptoms and go undiagnosed— could be as high as 86% prior to January 23, 2020. They estimated the transmission rate of undocumented infections as 55% of the rate for documented infections, and yet that undocumented infections contributed to 79% of documented cases. Another group of researchers found that the total contribution from the pre/asymptomatic population is more than that of symptomatic patients [26]. Future studies to estimate the asymptomatic ratio and the time when asymptomatic persons become infectious are needed to evaluate the effects of various control strategies.

The strength of this analysis is that it incorporates asymptomatic infections in the DP data that might not have been explored earlier. However, there are also limitations. First, due to inadequate data on the time course of infection cases among crew and passengers, assumption (d) (i) assumes a constant proportion, which may vary with time in practice. Second, the values of the parameter *r_p_* assumed in the present study may not be sufficiently large. Third, the assumptions (a)-(d) under chain-binomial models may not be sufficient to capture the complexity of the COVID-19 epidemics. Fuller data reporting is important for researchers to develop statistical methodology to help combat this pandemic.

Almost all of the passengers on DP were tested before they were evacuated. However, it is impractical to test everyone in the real world, especially for those asymptomatic cases. On DP, crew members continued to perform service unless they showed symptoms. This provides a parallel to people doing “essential work” in society and thus exempt from shelter-in-place rules. Our study suggests that if the asymptomatic ratio is high, the conventional quarantine might not be sufficient to reduce *R*_0_ to below 1, implying that a combination of preventive measures is needed to stop the spread of virus. When DP was docked in Taiwan for a 1-day tour on January 31, 2020, “big data analytics” was used to contain the spread of virus [27]. The low incidences in Taiwan strongly suggest that the virus can be contained with early and appropriate measures.

## Data Availability

This research is based on published data on the COVID-19 cases aboard the Diamond Princess, posted online by the National Institute of Infectious Disease, Japan.

https://www.niid.go.jp/niid/en/2019-ncov-e/9407-covid-dp-fe-01.html

https://www.niid.go.jp/niid/en/2019-ncov-e/9407-covid-dp-fe-02.html

